# “*I try not to go in order to hide my shame*”: A qualitative study exploring barriers and facilitators to mental health help-seeking among adolescents in Moshi Urban, Tanzania

**DOI:** 10.64898/2026.05.22.26353878

**Authors:** Ellyminius Mjuly, Irene Temba, Judith Kaale, Gloria Sechuma, William Nkenguye

## Abstract

**Background:** Adolescent mental health disorders represent a growing public health concern globally, with a substantial proportion of young people experiencing unmet mental health needs. Despite this burden, help-seeking behavior among adolescents remains low, particularly in low- and middle-income countries (LMICs), where structural, social, and cultural barriers persist. In Tanzania, limited context-specific evidence exists on factors influencing mental health help-seeking among adolescents, particularly within school settings.

**Methods:** A cross-sectional qualitative study was conducted among adolescents aged 15–19 years attending secondary schools in Moshi Urban, Kilimanjaro region, Tanzania, between April and May 2025. A total of 11 participants, including students, teachers, a school administrator, and a school healthcare provider, were recruited using convenience sampling. Data were collected using semi-structured questionnaires and focus group discussions, and audio-recorded for accuracy. Transcripts were analyzed using thematic analysis, following a systematic six-step approach. Codes were organized into subthemes and overarching themes.

**Results:** Three major themes emerged: facilitators, barriers, and suggested strategies for improving mental health help-seeking behavior. Key facilitators included the presence of school-based support systems, encouragement from trusted individuals (peers, parents, and teachers), perceived severity of mental health problems, and positive experiences from others. Major barriers included lack of trust and concerns about confidentiality, fear of information disclosure, stigma and fear of judgment, rigid school schedules, and poor teacher–student relationships. Participants highlighted the need for confidential, professionally led counselling services, increased mental health education, strengthened school-based programs, and improved access to mental health information as critical strategies to enhance help-seeking behavior.

**Conclusion:** Mental health help-seeking behavior among adolescents in Moshi Urban is influenced by a complex interplay of interpersonal, institutional, and individual factors. While supportive environments and social networks facilitate help-seeking, persistent barriers particularly related to trust, confidentiality, and stigma limit access to care. Strengthening school-based mental health services, improving mental health literacy, and ensuring confidential, youth-friendly support systems are essential to enhance help-seeking behavior and improve adolescent mental health outcomes in Tanzania and similar settings.

## Background

Mental health disorders represent a significant and growing public health burden globally, particularly among adolescents(1). Adolescence is a critical developmental period marked by rapid biological, psychological, and social transitions, during which individuals are especially vulnerable to mental health conditions(2). Globally, it is estimated that one in seven adolescents (approximately 14%) aged 10–19 years experience a mental disorder, accounting for nearly 15% of the global burden of disease in this age group(1–3). Depression, anxiety, and behavioral disorders are among the leading causes of illness and disability among adolescents, while suicide ranks as the third leading cause of death among young people aged 15–29 years(4). Importantly, nearly 75% of all mental health conditions begin before the age of 24, highlighting adolescence as a critical window for early intervention (5).

In Sub-Saharan Africa (SSA), the burden of adolescent mental health conditions is substantial and continues to rise(6,7). Approximately 13% of adolescents in SSA are estimated to have a diagnosable mental disorder, with higher prevalence reported for specific conditions such as depression (26.9%), anxiety (29.8%), and emotional or behavioral problems (up to 40.8%)(8). Furthermore, SSA has the fastest-growing adolescent population globally, with nearly a quarter of the population aged 10–19 years, amplifying the public health implications of unmet mental health needs in this region(8,9).

In Tanzania, emerging evidence suggests that adolescent mental health problems are both common and under-recognized(10). Studies among secondary school students indicate that approximately 29–33% of adolescents report at least one mental health problem, including symptoms of depression, anxiety, or emotional distress(5). Additional findings from national surveys show that 13.9% of adolescents have seriously considered suicide, 9.5% have made a suicide plan, and 11.5% have attempted suicide, underscoring the severity of the problem(5,11,12). Other studies in urban Tanzanian settings report depressive symptoms in approximately 9.4% of adolescents and suicidal expression in 13.4%, further highlighting the burden of mental health challenges among young people(13). Despite this, mental health services remain severely under-resourced, with approximately 0.06 psychiatrists and 0.01 psychologists per 100,000 population, reflecting a critical shortage of specialized care(14).

Despite the high burden, help-seeking behavior among adolescents remains markedly low. Help-seeking behavior involves recognizing mental health problems and actively seeking support from formal sources, such as healthcare providers, or informal sources, including family and peers(15). However, a substantial proportion of adolescents with mental health needs do not seek appropriate care. Globally, it is estimated that the majority of adolescents with mental health conditions remain untreated, particularly in low-resource settings(16,17). Barriers to help-seeking include stigma, low mental health literacy, fear of discrimination, concerns about confidentiality, and limited awareness or availability of services(18,19). In many African contexts, including Tanzania, cultural beliefs and reliance on traditional or informal care pathways further influence help-seeking behaviors(20).In Tanzania, these challenges are compounded by systemic and structural barriers. Mental health services are often centralized in urban referral hospitals, with limited integration into primary healthcare and school-based systems(21). Adolescents frequently rely on non-specialist providers, such as nurses or community health workers, who may have limited training in adolescent mental health care(22). Schools, which represent a key platform for early identification and intervention, often lack structured mental health programs, trained personnel, and referral mechanisms. At the same time, adolescents face multiple risk factors—including bullying, substance use, academic stress, and exposure to violence—which are strongly associated with increased mental health problems and poor help-seeking behavior(5).

Understanding the facilitators and barriers to mental health help-seeking behavior is therefore critical for improving adolescent mental health outcomes. Facilitators may include supportive family and peer networks, increased mental health awareness, availability of school-based services, and trust in healthcare providers. Conversely, barriers such as stigma, lack of confidentiality, negative perceptions of mental health services, and limited accessibility continue to hinder effective help-seeking(23). Importantly, these factors are highly context-specific and may vary across different socio-cultural and health system environments.

In Moshi Urban, Kilimanjaro region, there is limited context-specific evidence examining adolescent mental health help-seeking behavior, despite growing evidence of a substantial burden of mental health problems in this population. Without locally generated data, it remains difficult to design targeted interventions, strengthen school-based mental health services, or inform policy and programmatic decisions. Therefore, this study aims to assess the facilitators and barriers influencing mental health help-seeking behavior among adolescents attending secondary schools in Moshi Urban, Kilimanjaro. By generating context-specific evidence, the study seeks to inform the development of tailored interventions and contribute to strengthening adolescent mental health services in Tanzania and similar low-resource settings.

## Methods

### Study design

This study employed a cross-sectional qualitative design to explore facilitators and barriers influencing mental health help-seeking behavior among adolescents attending secondary schools in Moshi Urban, Kilimanjaro region, Tanzania. A qualitative approach was selected to enable an in-depth understanding of participants’ experiences, perceptions, and contextual factors shaping help-seeking behaviors. Data were collected over a defined period between April and May 2025.

### Study setting

The study was conducted in Moshi Urban, located in northern Tanzania within the Kilimanjaro region, which has an estimated population of 1,861,934, of whom approximately 23.8% reside in urban areas(24). Two secondary schools Old Moshi Secondary School and Mawenzi Secondary School were included in the study. Both schools offer ordinary and advanced level education, providing a diverse population of adolescents within an urban educational context.

### Ethical considerations

Ethical approval for the study was obtained from the KCMC University Research Ethics Committee, as well as relevant institutional authorities, including the Institute of Public Health and faculty leadership. Permission to conduct the study was also obtained from the headmasters of the participating schools. Written informed consent (and assent where applicable) was obtained from all participants. Confidentiality was maintained through the use of unique identifiers in place of names, and all data were securely stored. Participation was voluntary, and participants were informed of their right to withdraw from the study at any stage without providing a reason.

### Participants and eligibility criteria

The study population comprised adolescents aged 15–19 years enrolled in the selected secondary schools during the study period. In addition to students, key informants—including teachers, school administrators, and a school healthcare provider—were recruited to provide complementary perspectives on adolescent mental health help-seeking behavior. Participants were eligible if they were aged 15–19 years, enrolled in the selected schools, and willing to provide informed consent or assent. Adolescents who declined participation or were absent during the data collection period were excluded.

### Sampling and sample size

Participants were recruited using a convenience sampling approach based on availability, willingness to participate, and ability to communicate in English or Swahili. A total of 11 participants were included in the study. Sample size was guided by the principle of data saturation, defined as the point at which no new themes or insights emerge from the data. Saturation was achieved after six interviews; however, an additional five participants were included to confirm thematic redundancy and enhance analytical rigor.

### Data collection

Data were collected using semi-structured questionnaires supplemented by in-depth discussions. Separate tools were developed for students, school administrators, and the school healthcare provider to ensure that data captured reflected the perspectives of each group. The instruments included both open-ended and closed-ended questions designed to explore facilitators and barriers to mental health help-seeking behavior, as well as potential strategies for improvement. Data collection was conducted in both Swahili and English to ensure comprehension and inclusivity. All participants were provided with information about the study and gave written informed consent prior to participation, with assent obtained where applicable. Participants were informed of their right to withdraw from the study at any time without consequence.

Following questionnaire administration, focus group discussions consisting of approximately five students per group were conducted to deepen understanding of shared experiences and to triangulate findings across data sources. All discussions were audio-recorded with participants’ permission to ensure accuracy and completeness.

### Data processing and analysis

Audio recordings were transcribed verbatim and translated into English where necessary. Transcripts were organized according to participant groups (students, teachers, and administrators) and managed using QDA Miner software to facilitate systematic coding and data organization(25). Data were analyzed using thematic analysis, following a six-step approach: familiarization with the data; generation of initial codes; development of themes from coded data; review and refinement of themes; definition and naming of themes; and synthesis of findings into a coherent narrative. A total of 43 codes were identified and grouped into 13 subthemes, which were subsequently organized into three overarching themes: facilitators, barriers, and suggested strategies for improvement.

To enhance the trustworthiness of the findings, several strategies were employed. Credibility was strengthened through triangulation of data sources, including students, teachers, and administrators. Dependability was ensured through the use of a systematic coding framework and iterative discussions among four researchers involved in the analysis. Confirmability was supported by maintaining verbatim transcripts and an audit trail of analytical decisions, while transferability was enhanced through detailed description of the study context and participant characteristics.

## Results

### Social demographic characteristics of participants

A total of 11 participants, including students, teachers, school administrators, and a school healthcare provider, were included in the analysis(**Table 1**). Thematic analysis generated 43 codes, which were grouped into 13 subthemes and further organized into three overarching themes: (1) facilitators of mental health help-seeking behavior, (2) barriers to help-seeking, and (3) suggested strategies for improvement(**Table 2**).

**Table 1:**
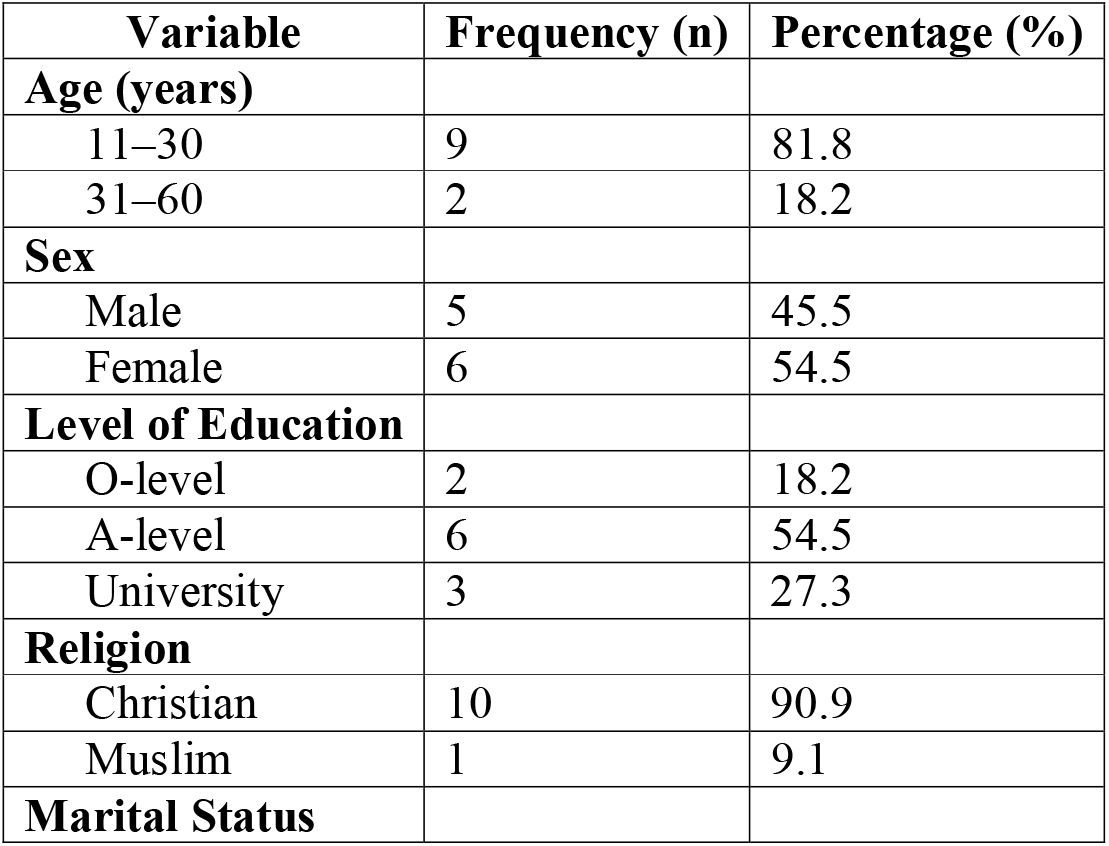
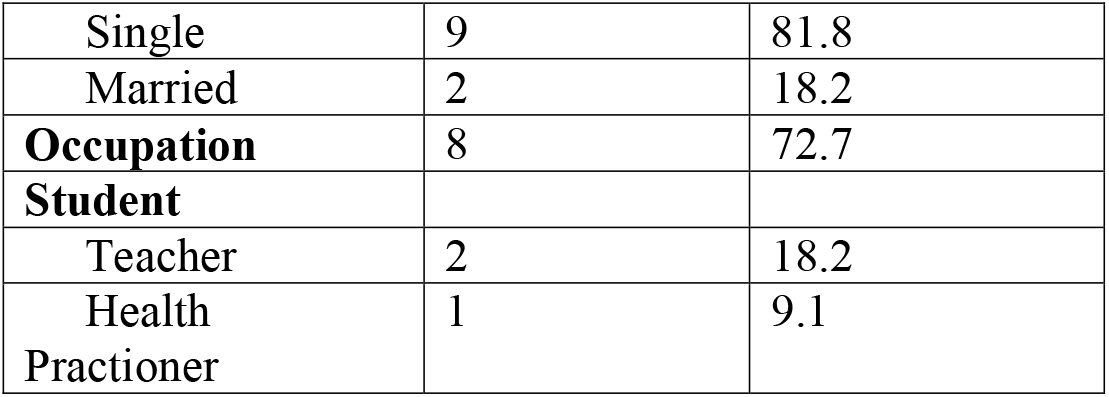
Social demographic characteristics of participants.

**Table 2:** Summary of Themes and Sub-themes on Mental Health Help-Seeking Behavior.

| Theme | Sub-themes |
| --- | --- |
| **Facilitators of Help-Seeking Behavior** | Supporting systems and school-based programs |
|  | Encouragement from parents, peers, and teachers |
|  | Perceived severity of mental health problems |
|  | Positive experiences from others |
| **Barriers to Help-Seeking Behavior** | Lack of trust and confidentiality |
|  | Fear of information disclosure |
|  | School-related constraints |
|  | Personal and emotional factors |
|  | Poor teacher–student relationships |
| **Strategies to Improve Help-Seeking Behavior** | Strengthening guidance and counselling services |
|  | Provision of mental health education |
|  | Enhancing school-based programs |
|  | Improving access to mental health information |

### Facilitators of mental health help-seeking behavior

Participants identified several factors that promoted mental health help-seeking among adolescents, including the presence of supportive school systems, interpersonal encouragement, perceived severity of mental health problems, and positive experiences from others.

### Supporting systems and school-based programs

School-based structures, including guidance and counselling units and student-led programs, were consistently reported as important facilitators. Participants highlighted the presence of formal and informal support systems such as guidance and counselling services, SHULE SALAMA, FEMA clubs, KISA clubs, and BINTI SALAMA initiatives, which provided safe spaces for students to express concerns and seek assistance. A school administrator noted:

> *“We have a Guidance and Counselling Unit staffed by two teachers who are available for consultation. Every teacher also plays a role as an advisor and is ready to support students whenever they face challenges” (School administrator, ID 07)*.

These structures were perceived as accessible and responsive, fostering an environment where students felt supported. Similarly, targeted programs addressing gender-specific challenges were reported to enhance help-seeking among female students:

> *“In our school we have an organization which states education for the right of female child; if we face any challenge, we consult the teacher in charge of the organization” (Student, ID 01)*.

### Encouragement from parents, peers, and teachers

Interpersonal relationships emerged as a critical facilitator of help-seeking behavior among adolescents. Participants emphasized that trust, familiarity, and emotional closeness with significant individuals—such as friends, parents, and teachers—created a supportive environment that enabled them to disclose personal challenges. These trusted relationships were often the first point of contact when adolescents experienced distress, as they provided a sense of safety, understanding, and reassurance. One participant explained:

> *“The one who can help is usually a close person, like a friend I trust, because it is easier to open up to someone who understands me; I can also involve my parent since they know me well, and sometimes even a teacher if I have built a good relationship with them, because they can guide me and give advice” (Student, ID 01)*.
>
> Participants further highlighted the importance of trust and emotional support in these relationships:
>
> *“When you have someone who listens to you without judging and shows they care, it becomes much easier to share your problems and seek help instead of keeping everything inside” (Student, ID 04)*.
>
> In addition, strong social connections were seen as protective and encouraging:
>
> *“Having supportive people around you gives you confidence that you are not alone, and it motivates you to speak out and look for solutions to your problems” (Student, ID 06)*.

### Perceived severity of mental health problems

Participants reported that help-seeking was often triggered when mental health challenges intensified and began to interfere with academic performance or daily functioning. Mild concerns were frequently managed independently, whereas worsening symptoms prompted adolescents to seek assistance. Many participants described a pattern in which help-seeking occurred only after problems became difficult to cope with, indicating a tendency toward delayed response rather than early intervention. One student explained:

> *“Most of us tend to seek help when the negative impact of a problem keeps getting worse, for example when someone starts failing exams or losing concentration in class, and at that point it becomes hard to manage alone, so we look for someone like a friend or teacher to help” (Student, ID 10)*.
>
> Participants further highlighted that early signs are often ignored or minimized:
>
> *“At the beginning, you may feel stressed or worried, but you think you can handle it yourself, so you don’t see the need to ask for help until it becomes too much” (Student, ID 04)*.
>
> In addition, some noted that worsening symptoms eventually force action:
>
> *“When the problem starts affecting your daily life or school performance, that is when you realize you need support because it is no longer something you can manage on your own” (Student, ID 06)*.

### Positive experiences from others

Observing or hearing about others’ successful experiences in addressing mental health challenges was reported as an important facilitator of help-seeking among adolescents. Participants explained that knowing someone who had faced similar difficulties and successfully sought help reduced fear, built trust in available support systems, and increased confidence in the possibility of recovery. Such shared experiences helped normalize mental health challenges and reassured students that they were not alone. One participant noted:

> *“If someone had the same problem and managed to solve it after seeking help, it gives others the courage to believe that their own situation can also improve, and it becomes easier to take that first step instead of staying silent” (Student, ID 09)*.
>
> Participants further emphasized that peer experiences played a key role in shaping attitudes toward help-seeking:
>
> *“When you hear a friend talk about how they got help and felt better, it makes you feel more confident that seeking support is useful and not something to be afraid of” (Student, ID 04)*.
>
> In addition, participants highlighted that visible positive outcomes reduced stigma and encouraged openness:
>
> *“Seeing others open up and improve shows that mental health problems can be managed, and that encourages more students to speak out and seek help” (Student, ID 06)*.

### Barriers to mental health help-seeking behavior

Despite the presence of facilitators, participants identified multiple barriers that hindered adolescents from seeking mental health support. These included lack of trust and confidentiality, fear of information disclosure, school-related constraints, personal and emotional factors, and poor teacher–student relationships.

### Lack of trust and confidentiality

The most frequently reported barrier was lack of trust, particularly concerns about confidentiality. Adolescents expressed fear that personal information shared during help-seeking could be disclosed to others, leading to stigma or embarrassment. One participant stated:

> *“I might speak out my problem then it becomes a chance for them to tell another person, which makes it difficult to seek help” (Student, ID 01)*.
>
> Similarly, another participant explained:
>
> *“Because of mistrust, the person may decide to keep things to themselves and find their own solution” (Student, ID 08)*.

### Fear of information disclosure

Closely related to issues of trust, fear of exposing personal issues was identified as a significant barrier to mental health help-seeking among adolescents. Participants expressed concerns about sharing sensitive information, particularly when it involved family matters or deeply personal experiences. Many feared that once disclosed, their problems could become widely known, leading to embarrassment, judgment, or social consequences. One student explained:

> *“If I say my problem, it feels like I am exposing it and making it public, and that makes me uncomfortable because I don’t know how others will react or who else might end up knowing about it” (Student, ID 01)*.
>
> Another participant emphasized uncertainty about confidentiality even among close peers:
>
> *“I can’t easily tell my friends about issues related to my family because I am not sure if they will keep it as a secret the way I would want, and that fear makes me keep things to myself” (Student, ID 03)*.
>
> Participants further noted that this fear of disclosure often leads to silence and avoidance of available support systems:
>
> *“Sometimes you have something really troubling you, but you choose not to talk about it because you are afraid it might spread beyond the person you trust” (Student, ID 05)*.

### School-related constraints

Structural barriers within the school environment, particularly rigid academic schedules, were identified as important factors limiting opportunities for mental health help-seeking.

Participants reported that tightly structured timetables left little or no time for students to access available support services, even when they were aware of them. This lack of flexibility made it difficult for students to seek help during school hours without disrupting their academic responsibilities. A school administrator explained:

> *“The timetable is very tight, from around 7:30 a*.*m. to 5:00 p*.*m. students are expected to be in class most of the time, so even if a student wants to seek help, there is hardly any free time to do so without missing lessons” (School administrator, ID 07)*.
>
> Students also emphasized how competing academic demands discouraged help-seeking:
>
> *“Sometimes you want to talk to someone or get help, but you keep postponing it because you are always in class or have assignments to complete, so the opportunity just passes” (Student, ID 04)*.
>
> In addition, participants noted that the absence of designated time for counselling or support services further limited utilization:
>
> *“If there was a specific time set aside for counselling or support, more students would be able to attend without worrying about missing classes” (Student, ID 06)*.

### Personal and emotional factors

Internal factors, including fear of judgment and feelings of shame, were identified as significant barriers to mental health help-seeking among adolescents. Participants reported that concerns about how others—particularly teachers and peers—might perceive them often discouraged them from seeking support. This fear was closely linked to anticipated stigma, negative labeling, and misunderstanding of their problems, leading many students to suppress their emotions rather than disclose them. One student explained:

> *“I feel worried about how the teacher will perceive me when I ask for help, because sometimes you think they might judge you, misunderstand your situation, or see you as weak instead of trying to support you” (Student, ID 10)*.
>
> Another participant highlighted how shame and fear of exposure further limited help-seeking:
>
> *“I try not to go in order to hide my shame, because some problems feel too personal or difficult for others to understand, and you worry that once you say it out loud, people might look at you differently or treat you in another way” (Student, ID 01)*.
>
> Participants also noted that these internal struggles often lead to isolation and delayed care-seeking:
>
> *“Sometimes you keep everything inside because you are not sure how others will react, and that makes it harder to ask for help even when you really need it” (Student, ID 05)*.

### Poor teacher–student relationships

The quality of relationships between students and teachers was identified as a key factor influencing willingness to seek help for mental health concerns. Participants reported that lack of closeness, approachability, and trust in teachers significantly limited engagement with available support systems. When students perceived teachers as distant, judgmental, or unapproachable, they were less likely to disclose personal challenges or seek guidance, even when support services were available. One student explained:

> *“If the teacher is close enough and approachable, students feel comfortable to consult and share their problems, but if there is no trust or connection, most of us prefer to keep things to ourselves instead of seeking help” (Student, ID 02)*.
>
> Participants further emphasized that supportive and understanding teacher–student relationships could foster a safe environment for help-seeking:
>
> *“When a teacher listens without judging and shows understanding, it becomes easier for students to open up and ask for help, but if they seem strict or dismissive, students will avoid them” (Student, ID 05)*.
>
> In addition, some students highlighted that fear of negative reactions or lack of empathy from teachers discouraged disclosure:
>
> *“Sometimes we are afraid that teachers may not understand our situation or may judge us, so we choose not to speak about our problems at all” (Student, ID 03)*.

### Suggested strategies for improving help-seeking behavior

Participants proposed several strategies to enhance mental health help-seeking among adolescents, focusing on strengthening counselling services, increasing mental health education, improving school programs, and enhancing access to information.

### Strengthening guidance and counselling services

Participants emphasized the need for dedicated, confidential, and professionally led counselling services within schools as a critical strategy to improve mental health help-seeking among adolescents. They highlighted that while some support structures exist, the absence of specialized personnel and private spaces limits their effectiveness.

Recommendations included establishing well-equipped and private counselling rooms, employing trained mental health professionals, ensuring gender-sensitive staffing, and implementing structured follow-up mechanisms to support students over time. One teacher underscored this need by stating:

> *“There should be a specific room for counselling that ensures privacy, and it should be managed by trained professionals who understand mental health issues, because students are more likely to open up when they feel safe and are confident that the person helping them has the right expertise” (Teacher, ID 06)*.
>
> Students also emphasized the importance of confidentiality and trust in professional services:
>
> *“I trust an expert because they are trained to handle personal issues and keep them confidential, just like doctors do with their patients, so it becomes easier to share my problems without worrying that my secrets will be exposed or judged by others” (Student, ID 03)*.
>
> Participants further noted that consistent follow-up and accessibility of counselling services are essential for sustained support:
>
> “It is important that counselling is not just a one-time conversation, but that there is follow-up to check on the student’s progress, so they feel supported throughout their situation” (Student, ID 05).

### Provision of mental health education

Participants emphasized the importance of increasing mental health education within schools as a key strategy to improve awareness, reduce stigma, and promote early help-seeking behavior. Many noted that limited understanding of mental health conditions contributes to misconceptions, fear, and delayed care-seeking among students. Enhancing mental health literacy was therefore viewed as essential in normalizing conversations around psychological well-being and equipping students with the confidence to seek help. A school administrator highlighted this need, stating:

> *“More education should be provided on mental health issues, and experts should be invited to schools to clearly explain the different types of mental health problems, their causes, signs, and how they can be managed, so that students can better understand what they are experiencing and know where and how to seek help without fear, confusion, or stigma” (School administrator, ID 07)*.
>
> Students further emphasized that structured education sessions could create a safe environment for open discussion and reduce stigma:
>
> *“If we are taught about mental health in a detailed and practical way, including how to recognize symptoms and cope with challenges, it helps us understand that these problems are normal and treatable, and it becomes easier to talk about them openly instead of keeping everything to ourselves” (Student, ID 04)*.
>
> Participants also suggested integrating mental health education into routine school activities to promote early recognition and response:
>
> *“When mental health is taught regularly in school, students can notice early changes in their thoughts, emotions, or behavior, support their friends who may be struggling, and seek help early before the situation becomes serious or starts affecting their academic performance and daily life” (Student, ID 06)*.

### Enhancing school-based programs

Strengthening existing programs and introducing new initiatives were identified as important strategies to improve mental health help-seeking among adolescents. Participants emphasized that while several school-based platforms already exist, their effectiveness depends on consistent monitoring, active engagement, and proper coordination. There was a strong call for ensuring that these programs are not only established but also remain functional, visible, and accessible to students who need support. One student highlighted this by stating:

> *“Clubs should be followed up to make sure they are actively operating, not just existing in name, so that students can consistently participate, share their challenges, and receive support in a structured and safe environment” (Student, ID 02)*.
>
> Participants further noted that strengthening these programs requires committed leadership, regular supervision, and clear accountability mechanisms:
>
> *“If these programs are well monitored and supported by teachers, they can become a reliable space where students feel comfortable discussing their problems and learning from each other” (Student, ID 05)*.
>
> In addition, expanding the scope of school-based initiatives to include more targeted mental health activities was seen as beneficial:
>
> *“Schools should introduce more programs that focus specifically on mental health, with regular sessions, discussions, and activities that encourage students to speak openly and support one another” (Student, ID 03)*.

### Improving access to mental health information

Participants highlighted the need to increase access to mental health resources within schools as an important strategy to support help-seeking behavior among adolescents. They noted that limited availability of reliable information often leaves students uncertain about how to understand or manage their mental health challenges. Expanding access to educational materials was therefore seen as essential in promoting awareness, improving knowledge, and empowering students to make informed decisions about seeking help. One student emphasized this by stating:

> *“The school should purchase psychology books and other mental health materials and prepare a proper study space where students can read, learn, and understand different mental health conditions, their causes, and how they can be managed without feeling lost or confused” (Student, ID 08)*.
>
> Participants further suggested that making such information easily accessible within the school environment could encourage independent learning and reduce stigma:
>
> *“If mental health information is available in places like the library or notice boards, students can learn privately at their own pace, which helps them understand their problems better and reduces fear of being judged” (Student, ID 05)*.
>
> In addition, participants emphasized the importance of creating supportive and conducive environments that promote learning and reflection:
>
> *“A quiet and comfortable space where students can read, reflect, and access information about mental health can help them recognize their struggles early and feel more confident about seeking help” (Student, ID 03)*.

## Discussion

This study explored facilitators and barriers influencing mental health help-seeking behavior among adolescents attending secondary schools in Moshi Urban, Tanzania. The findings highlight a complex interplay of individual, interpersonal, and structural factors shaping adolescents’ decisions to seek support. While supportive school systems and social networks facilitated help-seeking, persistent concerns around trust, confidentiality, stigma, and institutional constraints emerged as major barriers.

Consistent with previous studies conducted in sub-Saharan Africa and other low-and middle-income countries (LMICs), this study found that interpersonal relationships particularly with friends, parents, and teachers play a central role in facilitating help-seeking behavior.

Adolescents were more likely to seek help from individuals they trusted and felt emotionally connected to. Similar findings have been reported in studies from Kenya(26), Uganda(27), and South Africa(28), where peer and family support were identified as key entry points into mental health care. These findings align with social support theories, which suggest that trusted relationships reduce psychological distress and increase openness to seeking assistance.

School-based structures, including guidance and counselling units and student-led programs, were also identified as important facilitators. This is consistent with global evidence emphasizing schools as critical platforms for early identification and intervention in adolescent mental health. However, despite the presence of these systems, their effectiveness appeared to be limited by structural and relational factors, suggesting that availability alone is insufficient without quality, accessibility, and trust(29).

A key finding of this study is that help-seeking among adolescents is often reactive rather than preventive, with many participants reporting that they only seek support when problems become severe and begin to affect academic performance. This pattern has been widely documented in adolescent populations globally and reflects low mental health literacy, delayed recognition of symptoms, and normalization of distress until it becomes overwhelming(30). This finding underscores the need for early intervention strategies and mental health education within school settings.

The most prominent barriers identified were lack of trust and concerns about confidentiality. Adolescents expressed strong fears that personal information disclosed during help-seeking would be shared with others, leading to stigma, embarrassment, or social consequences. Similar concerns have been consistently reported across studies in both high-income and low-resource settings, where perceived breaches of confidentiality significantly reduce willingness to seek help(31). In the present study, this fear was not only directed toward formal support systems but also toward peers, highlighting the pervasive nature of mistrust in adolescent social environments.

Fear of judgment and stigma also emerged as critical barriers, with adolescents expressing concerns about how they would be perceived by teachers and others if they sought help(32). These findings are consistent with global literature demonstrating that stigma remains one of the most significant deterrents to mental health service utilization among young people. Internalized stigma, including feelings of shame and perceived weakness, further compounds these barriers and contributes to avoidance of help-seeking.

Structural barriers within the school environment, particularly rigid academic schedules, were also identified as limiting access to mental health support. Even where services were available, time constraints reduced their utilization(33). This finding aligns with evidence from school-based studies indicating that competing academic demands often take precedence over mental health needs, particularly in resource-constrained educational systems.

The quality of teacher–student relationships was another important determinant of help-seeking behavior. Adolescents were more likely to seek support from teachers they perceived as approachable and trustworthy. Conversely, distant or formal relationships discouraged engagement with available support systems(34). This highlights the importance of relational dynamics in shaping access to care, particularly in school settings where teachers often serve as first points of contact.

Participants proposed several strategies to improve help-seeking behavior, including strengthening guidance and counselling services, increasing mental health education, expanding school-based programs, and improving access to mental health information. Notably, adolescents emphasized the need for professional, confidential, and dedicated mental health services, rather than relying solely on teachers(35). This reflects a broader recognition of the importance of specialized care and aligns with global recommendations advocating for integration of trained mental health professionals within school systems.

### Strengths and limitations

This study has several strengths. First, the use of a qualitative design allowed for an in-depth exploration of adolescents’ experiences and perceptions, providing rich, context-specific insights into mental health help-seeking behavior. Second, the inclusion of multiple participant groups—students, teachers, and school administrators—enabled triangulation of perspectives, enhancing the credibility and depth of the findings. Third, the study applied a systematic thematic analysis approach, with collaborative coding and iterative discussions among multiple researchers, strengthening the reliability of the analysis.

However, the study also has limitations. The use of a convenience sampling strategy and a relatively small sample size may limit the transferability of the findings to other settings. Additionally, the study was conducted in two schools within an urban area, which may not reflect the experiences of adolescents in rural settings or other regions of Tanzania. Social desirability bias may have influenced participants’ responses, particularly given the sensitivity of mental health topics. Furthermore, reliance on self-reported data may introduce recall or reporting bias. Despite these limitations, the study provides valuable insights into an under-researched area and highlights important directions for future research and intervention.

## Conclusion

This study highlights that mental health help-seeking behavior among adolescents in Moshi Urban is shaped by a complex interaction of supportive systems, social relationships, individual perceptions, and structural constraints. While school-based programs and interpersonal support networks facilitate help-seeking, significant barriers—including lack of trust, concerns about confidentiality, stigma, and limited access within school environments—continue to hinder effective utilization of mental health services.

Addressing these barriers requires a multifaceted approach, including strengthening confidential and professional counselling services within schools, enhancing mental health literacy among adolescents, fostering supportive teacher–student relationships, and integrating mental health into school policies and programs. Interventions should be context-specific and adolescent-centered, taking into account the social and cultural dynamics influencing help-seeking behavior. Strengthening school-based mental health systems has the potential to improve early identification, increase service utilization, and ultimately enhance mental health outcomes among adolescents in Tanzania and similar resource-limited settings.

## Data Availability Statement

Deidentified data is available upong request

